# Measuring health literacy from the communal perspective: initial testing of the Information and Support for Health Actions Questionnaire (ISHAQ)

**DOI:** 10.64898/2026.08.04.26359664

**Authors:** Christina Cheng, Roy Batterham, Gerald R Elsworth, Saichon Kloyiam, Charay Vicathai, Napaporn Wanitkun, Melanie Hawkins, Richard H Osborne

## Abstract

Measuring health literacy is important in addressing health inequity. However, current measures are developed from the individualistic perspective that values personal autonomy and choice. Applying such measures to people from a communal culture that values collective actions may lead to biased responses. To address this gap, a health literacy measure drawing on the communal perspective was developed in Thailand. With the aim to also develop an equitable measure, a grounded approach including strategies to include people with special needs such as people with chronic illness or physical disabilities, blind people and deaf people, was used. Concept mapping workshops were conducted, involving 254 participants including general community members, people with special needs, health professionals and policymakers. The result was a draft questionnaire of 17 hypothesized scales. Psychometric testing involving a survey of 2,228 participants resulted in a 14-scale questionnaire, the Information and Support for Health Actions Questionnaire (ISHAQ). This paper reports on the psychometric testing of the 14-scale ISHAQ and its supplementary scales for people with special needs. Item difficulty, scale reliability, one-factor confirmatory factor analysis using robust maximum likelihood estimator, and measurement invariance across groups with special needs using the alignment method with Bayesian estimation, were evaluated. A total of 2,262 respondents participated in the survey. Six scales achieved excellent model fit while one with reasonable fit and seven scales achieved reasonable to excellent fit following modifications. Supplementary scales also achieved reasonable to excellent model fit. Reliability for all scales were acceptable to good. Measurement invariance was confirmed for eight scales. With strong validity evidence, the ISHAQ is translated into English and ready for implementation. With the potential to be applied in different settings given cultures exist in a continuum, the ISHAQ can be used for health literacy needs assessment to support intervention development to improve health outcomes and equity.

## Introduction

### Measuring health literacy

Health literacy is intrinsically linked to health equity [1, 2]. Measuring health literacy is an important step to address issues of health inequity [3]. However, the development of a measure is often influenced by the context, worldview, perspective and construct definitions of the developers [4]. When a measure is applied to people coming from another context with different worldview or perspective, responses may become biased and misrepresented [5], leading to the potential of unfair measurement [5], which may exacerbate health inequity.

Health literacy is a concept first coined in 1974 in the United States. It started out as a component of health education relating to people’s ability in reading and writing [6]. Early definitions centred around how an individual needed to understand information in order to maintain good health [2]. Hence, health literacy measures developed in accordance with the early definitions were tools focused on reading, writing and numeracy.

By the 2000s, the definitions of health literacy begun to shift from solely about the individual to the demands of the systems and even linked to the societal level of a group or community [7]. Such thinking uncovered the limitations of measures that focused only on reading and writing [2]. It has also been pointed out that higher level of reading and writing skills is not a guarantee that people will respond positively to health education [8] and literacy screening may even run the risks of shame and stigmatisation [9].

Health literacy is now generally recognised as a multidimensional concept involving settings [10] as well as social and cultural contexts [11–13], and multidimensional measures of health literacy have been developed. A 2022 systematic review of health literacy measures identified a number of multidimensional measures [14], with the European Health Literacy Survey Questionnaire (HLS-EU-Q) [15] and the Health Literacy Questionnaire (HLQ) [16] being the two most commonly used.

As a concept first started in the United States, development of further theories and research mostly take place in Australia, Europe and North America, where individual autonomy and choice over health and wellbeing frames the thinking behind the development of health literacy measures [17]. However, increasing experiences of working with people from indigenous and minority communities as well as other countries around the world has broadened the thinking of health literacy. In fact, ‘for many people in most parts of the world, health decisions and actions are part of family, community or cultural processes, practices, beliefs or religious teachings’ [17].

The most recent definition of the health literacy of an individual from the World Health Organization (WHO) takes a globally relevant perspective and refers it as ‘people’s knowledge, confidence and comfort – which accumulate through daily activities and social interactions and across generations – to *access, understand, appraise, remember* and *use* information about health and health care, for the health and wellbeing of themselves and those around them’ [17]. Hence, applying a health literacy measure, framed in an individualist perspective that values individual autonomy and personal choice, to people coming from the communal culture where health is often a collective action and decision, has the potential to lead to biased responses, resulting in misinterpretation of data [4]. Bhakuni and Abimbola pointed out that when insufficient attention is given to the possibility that interpretive tools, conceptual or knowledge frameworks may not apply to certain groups, epistemic injustice occurs [18].

Among the multidimensional tools, the nine-scale HLQ may be one of the first health literacy measures to include a social dimension. It was developed using a grounded approach to identify health literacy domains based on extensive consultation with community members and clinical staff in Australia and international health experts [16]. The HLQ has been translated into 40 languages since its inception in 2013. Every care has been taken when the HLQ is translated into a different language and adapt into a different culture to ensure construct equivalence. The tool has been successfully used in over 90 countries to identify the health literacy strengths and challenges of survey participants in various contexts and settings [19–23] and support intervention development with promising positive outcomes [24, 25]. Yet, the HLQ is still developed mostly in Australia based on a predominantly individualistic perspective. A 2022 systematic review of health literacy measures reported on a number of instruments developed in countries dominated by communal culture such as China, Japan, Korean, Iran and Zambia [14]. However, all the tools were developed based on the western concept of health literacy with most of them still focus on functional health literacy, i.e., about basic skills in reading and writing health information or on knowledge of health facts [8].

In view of the lack of a health literacy measure capturing the communal culture, a collaboration between the HLQ developers, an Australian team, with the Thai Ministry for Public Health, the Health System Research Institute and Mahidol University initiated the development of a tool based on the perspective of the communal culture of Thailand in 2008.

### Developing an equitable measure from the communal perspective

The Information and Support for Health Actions Questionnaire (ISHAQ) was developed, in Thai language, with specific strategies applied to build a tool with the capacity for equitable measurement. The tool has been used as simply ‘a Thai health literacy questionnaire’ in Thailand [26] and ethical approval for the development process was granted under that name. ISHAQ was chosen for internationalisation. From the beginning, a focus on grounded and participatory development was emphasised. The process started with an extensive process of consultation involving 31 concept mapping groups across all regions of Thailand. Participants included people from the general population (people self-reported as being well), people with chronic illness, people with physical disabilities, blind people, deaf people, health service providers and health policy makers. New processes for concept mapping were specifically developed to support the participation of the deaf and blind groups.

At each concept mapping workshop, participants were first asked about what they needed to access and apply health information to take care of their own health. Then, they were asked to think the same for other people including their family, close friends and people in the community. From the answers of the 254 workshop participants, statements were analysed using cluster analysis to identify a range of common themes across the groups, from high level themes to more detailed and specific themes. See the original measurement model for the general population in S1 Figure. The integrated themes, and the statements from the groups associated with these themes, were used to develop a draft questionnaire including 17 hypothesized scales and 105 draft items, plus supplementary scales focus on people with chronic illness, people with physical disabilities, blind people and deaf people.

An extensive process of cognitive testing across different regions in Thailand was then undertaken. The testing involved each of the groups listed above, with the purpose to ensure that the draft questions were equally understandable and answerable across the groups. In addition, different administration formats were also tested, including in paper format, face-to-face interview and in sign language presented in the form of a digital video disc.

Based on the results from the cognitive testing, a total of 16 scales (nine Health knowledge and capabilities scales, six Health actions scales and one Enabling scale) with 88 items for the general population, supplementary scales for people with chronic illness (three scales), physical disability (two scales), blindness (three scales) and deafness (three scales) were developed. An initial validity testing was undertaken in 2012 with a sample size of 2,282 survey respondents including people from the general population as well as people with special needs including people with chronic illness, physical disabilities, blind people and deaf people. Respondents were asked to rate each item on a 0-10 scale anchored at the ends of the scale by ‘strongest possible disagreement’ to ‘strongest possible agreement’. Item selection analyses and preliminary scale validation were conducted in the tradition of classical test theory and latent variable analysis. Given the items were set a priori to each scale, confirmatory factor analysis (CFA) was conducted.

Model fit of the 16 one-factor models was judged to be excellent fit for one, close fit for six scales, reasonable fit for two scales and unsatisfactory fit for six scales while one scale with only three items could not be estimated. Five scales achieved excellent fit after model modification with one other led to reasonable fit. However, two scales with unsatisfactory fit could not be improved. Factor loadings were generally acceptable for most items, with the loading of 37 items ≥ 0.7 while 13 items were <0.5. Factor analysis-based composite reliability was acceptable (≥ 0.70) for 14 of the 16 scales while Cronbach’s alpha for the 3-item scale was 0.60. See S2 Table for results of this study.

Informed by these results, the ISHAQ was revised, and a second validity testing study was conducted. This paper presents the validity evidence on the internal structure and reliability generated from the psychometric analyses of the revised ISHAQ in Thai language.

## Materials and Methods

Based on the initial development and validity testing results, a further selection of scales and items was updated by the research team. A second validation study with a separate sample of respondents was then conducted. Using a cross-sectional design, data were collected in 2013.

### The Information and Support for Health Actions Questionnaire (ISHAQ)

By reviewing the initial validity testing results, the ISHAQ was revised to a questionnaire with 14 scales including eight Health knowledge and capabilities scales, five Health actions scales and one Enabling scale with 68 items for the general population, with supplementary scales for people with chronic illness (two scales), physical disability (two scales), blindness (three scales) and deafness (three scales). Some of the scale names and items were also refined (S2 Table). See Table 1 for the scales and number of items for each scale. The questionnaire continued to retain the 11 response options of a 0-10 scale from ‘strongest possible disagreement’ to ‘strongest possible agreement’.

**Table 1.** The scales and number of items of the Information and Support for Health Actions Questionnaire (ISHAQ) in English

| Scale | Number of items |
| --- | --- |
| **Health knowledge and capabilities scales** |  |
| 1. Receiving knowledge about basic rights | 4 |
| 2. Supporting health in the community | 6 |
| 3. Ability in receiving health services | 4 |
| 4. Communication skills to get what you want from health professionals | 5 |
| 5. Family health | 5 |
| 6. Ability to find suitable health information | 7 |
| 7. Assessing the believability of health information | 4 |
| 8. Accepting responsibility for health | 5 |
| **Health actions scales** |  |
| 9. Ability to choose healthy food | 4 |
| 10. Exercise for health | 5 |
| 11. Managing stress | 4 |
| 12. Using medicines | 5 |
| 13. Using herbs and supplements | 5 |
| **Enabling scale** |  |
| 14. Travelling barriers and ability | 5 |
| **Supplementary scales for people with special needs** |  |
| **Chronic illness group** |  |
| C1. Exchange experience and knowledge with other patients | 5 |
| C2. Self-monitoring | 8 |
| **Physical disability group** |  |
| P1. Access to health services in hospitals* | 5 |
| P2. Equipment^ | 4 |
| **Blind group** |  |
| B1. Access to health services in hospitals* | 5 |
| B2. Equipment^ | 4 |
| B3. Access and use | 4 |
| **Deaf group** |  |
| D1. Access to health services in hospitals* | 5 |
| D2. Equipment^ | 4 |
| D3. Access and use of interpreters | 4 |

\* or ^ Common scales across the Physical disability, Blind and Deaf groups.

### Participants

People aged 18 years old or over and could understand Thai were eligible to participate in the survey. Additional criteria were added for special needs group scales relevant to the specific groups. While each participant received a paper copy of the survey, an interviewer read out the questions for the participants to answer to ensure people of all literacy levels could participate. For deaf people, a video using sign language was played and the participants answered each question along with the video.

### Recruitment

Data were collected from five regions across urban and rural Thailand including Northern, Northeastern, Western, Central, Eastern and Southern Thailand using tailored sampling and recruitment strategies for the different groups to ensure broad representation. For the general population, stratified random sampling was used. Target provinces were divided into 25–50 sub-units, from which survey locations were randomly selected. Additional units were randomly drawn when needed to achieve sample targets. Researchers also applied quota controls for gender and age (18–60 years and 61 years or over) based on demographic data from the National Statistical Office’s 2012 fourth quarter labour survey.

For people with chronic illness, a mixed approach was adopted to reduce reliance on hospital-based sampling. Twenty-five percent of participants were recruited from provincial hospitals during service visits, while 75% were recruited through community field surveys, supported by coordination with hospitals and community leaders.

People with physical disability and blind people were recruited using the Health Systems Research Institute (HSRI) disability registries and local community networks. Researchers worked with local agencies and community leaders to identify and access eligible individuals. Where registry-based recruitment was insufficient, additional participants were recruited through community outreach, including visits timed to coincide with community gatherings. Recruitment of blind people in southern provinces required expansion to Phuket due to insufficient numbers in Songkhla and Surat Thani. For deaf people, direct field data collection was conducted by the HSRI.

## Statistical analysis

Descriptive statistics were used for the demographic data while the psychometric analyses followed the tradition of classical test theory. All analyses were conducted using SPSS Version 22 [27] and Mplus Versions 7 and 8 [28]. For conducting CFA, there is no consensus on the best way to estimate sample size. Recommendations may range from 100 to 400 or a minimum of 200 or 5 to 20 respondents per observed variable [29]. For the handling of missing data, the default estimation of full information maximum likelihood (FIML) in Mplus was used. This estimation allows for the use of all available data on all items.

Given the purpose of developing an equitable measure and the real possibility that the population heterogeneity generated by the sampling of this study would make pooling data for psychometric analysis across the five selected groups problematic, this study considered different strategies and decided to use the general population group as the reference group. Classical item analysis and confirmatory factor analyses were conducted using this reference group and options for scale improvement, if needed, were generated from the results of these analyses. These options of improvement were tested based on the data from the general population group, followed by the investigation of measurement invariance across the data from all five groups. The final decision for item selection or revision was based on the results as well as expert judgement of the content validity of relevant items.

## Psychometric properties

The analyses first assessed the psychometric properties of items hypothesised to measure a single homogeneous scale using data from the reference group, i.e., the general population. Four analyses were involved: a) item difficulty; b) scale reliability; c) scale reliability if the item were to be deleted; d) one-factor CFA model fitted to the data for the proposed scale.

With the 11-response option (0-10), the item difficulty index was dichotomous and calculated as the proportion responding 6–10 on the scale as against the proportion responding 0–5. It was scaled in the direction of item ‘easiness’ – the higher the score, the greater the proportion of people responding 6–10, that is towards the ‘strongest possible agreement’ pole of the response continuum. For scale reliability, it is well known that the widely used index of Cronbach’s alpha can be a biased estimate of reliability when the scale components are not tau-equivalent, i.e., do not have equal factor loadings [30] and item errors may be correlated [31], which are likely in the case of the ISHAQ. Hence, Raykov’s method of composite reliability which linked directly to the CFA of the hypothesised scales was used [32, 33], providing a coherent program of item analysis and collection of statistical evidence of scale consistency and reliability. A reliability of ≥ 0.70 and < 0.95 was considered acceptable to good [34].

For the one-factor CFA for the proposed scale, the purpose was to locate a model that yielded a set of items that demonstrated unidimensionality. This is essential when these constructs are to be used for needs assessment and/or program evaluation where unambiguous construct measurement is essential [35, 36]. Initial assessment of the data found that, with the response continuum containing 11 response options, many distributions were noticeably left skewed and leptokurtic, particularly in the reference group, suggesting potential ceiling effects. Therefore, the CFA models were fitted to the data using the robust maximum likelihood (MLR) estimator available in Mplus that provides fit indices and standard errors that are robust to the violation of normality. Besides, ordinal approaches cannot be used for over 10 response options in Mplus. Based on the Mplus outputs, standardized factor loadings and fit statistics were examined. Factor loading is generally considered acceptable if it is over 0.50 [37] but a threshold value of over 0.60 can support an acceptable construct reliability for scales with 3 or more items [38]. For the fit indices, the threshold value for the probability for an acceptable value of the chi-square test of the ‘exact fit’ of the hypothesized model to the data was set at the p-value of ≥0.01. However, since Chi-square tests of exact model fit are directly sensitive to sample size, they were not taken as the single piece of evidence for rejecting a model if Chi-square was statistically significant. Rather, the tests of ‘close fit’ were also considered, including Comparative fit index (CFI) ≥0.95, Tucker-Lewis fit index (TLI) ≥0.95, Root mean square error of approximation (RMSEA) ≤0.06 and standardized root mean residual (SRMR) ≤0.08 while a value of RMSEA ≤0.08 indicates a ‘reasonable fit’ [39].

To further determine model fit, the modification index (MI) and standardised expected parameter change (SEPC) were examined to identify potential correlated residuals in the one-factor models. A MI ≥ 3.84 is defined as statistically significant and, when associated with an SEPC of ≥ 0.2, it is taken to indicate the need to include an additional parameter in the model following careful consideration of theoretical plausibility [40, 41]. Should the correlated residuals point to the possibility of a multi-factor solution within a scale, exploratory structural equation modelling (ESEM) was used. ESEM is a less restrictive approach than CFA and uses target rotation to model data in a confirmatory way by allowing cross-loadings between items. However, the cross-loadings are constrained to close to zero to prevent increased parameters or misleading model fit. The ESEM approach may provide a better understanding of the associations of the items to the hypothesised construct [42, 43].

### Measurement invariance

When group comparisons are informed by multi-item composite scales, it is important to establish measurement invariance across these groups. This includes configural invariance (same factor structure), metric invariance (factor loadings being equivalent) and scalar invariance (item intercepts being equivalent) [44].

Traditionally, evaluating measurement invariance was tested by multiple-group CFA which presented many challenges such as many necessary model modifications [45, 46].

An alternative method, the alignment method or alignment optimisation, was proposed by Asparouhov and Muthén in 2013. They have also programmed this method in the Mplus software. The method is designed to enable the unbiased estimation of the factor means of multiple groups “without requiring exact measurement invariance” and is based on the configural invariance model. After fitting the configural model, the alignment optimization approach then uses a simplicity criterion (analogous to that used in factor rotation to simple structure) to locate the most optimal pattern of measurement invariance across groups. The chosen simplicity criterion is a loss function that is minimised when there are many approximately invariant measurement parameters (item factor loadings and item intercepts) and a small number of large non-invariant measurement parameters [45].

The program output provides information on the fit of the configural models (fit for All groups combined, followed by each of the five groups, i.e., General, Chronic illness, Physical disability, Blind and Deaf groups) and the pattern of optimised invariance/non-invariance of each measured variable across each of the five groups, in addition to significantly different factor means based on the optimised invariance pattern. The alignment method can be used with both maximum likelihood and Bayesian estimation. As initial assessment of the data showed strong non-normal distributions for many of the scales, therefore, the Bayesian approach, which does not rely on large sample theory and gives better performance in smaller samples [47], was used. To evaluate model fit, the posterior predictive p (PPP) value and 95% credibility interval for the difference between the observed and replicated chi-square values were examined. A PPP value of around 0.5 and a value of 0 falling close to the middle of the 95% credibility interval indicates excellent fit while a low PPP value and positive 95% credibility interval denotes poor fit [47]. Based on Monte Carlo simulations, Asparouhov and Muthén suggested that a limit of 25% of non-invariance (or 75% of total invariance) parameters may be safe for trustworthy alignment results [48].

### Ethics

This study was approved by Mahidol University Institutional Review Board (MU-IRB). The ethical approval number of the first study presented in the Introduction is MU-IRB2012/062.2903 and that of this study is MU-IRB 2011/134.2211. Oral consent was obtained, as approved by MU-IRB, and documented by the interviewer who conducted the survey interview. As mentioned above, face-to-face interviews, hence oral consents, were conducted to ensure people of all literacy levels could participate.

## Results

### Participant characteristics

The survey for the ISHAQ psychometric testing was administered to 2,262 respondents across five regions of Thailand. Close to a quarter of the participants lived in the Eastern region (24.8%), North region (23.5%) and Northeast region (24.3%), while 50.8% of them lived in urban areas. There were 57.0% female and 43.0% male respondents, with an average age of 51.7 (standard deviation, SD=17.8). About two-thirds of participants (67.5%) only had primary or below education. Most of the participants (90.5%) lived with their families, with an average number of household members being 3.4 (SD=2.0). See Table 2.

**Table 2.**
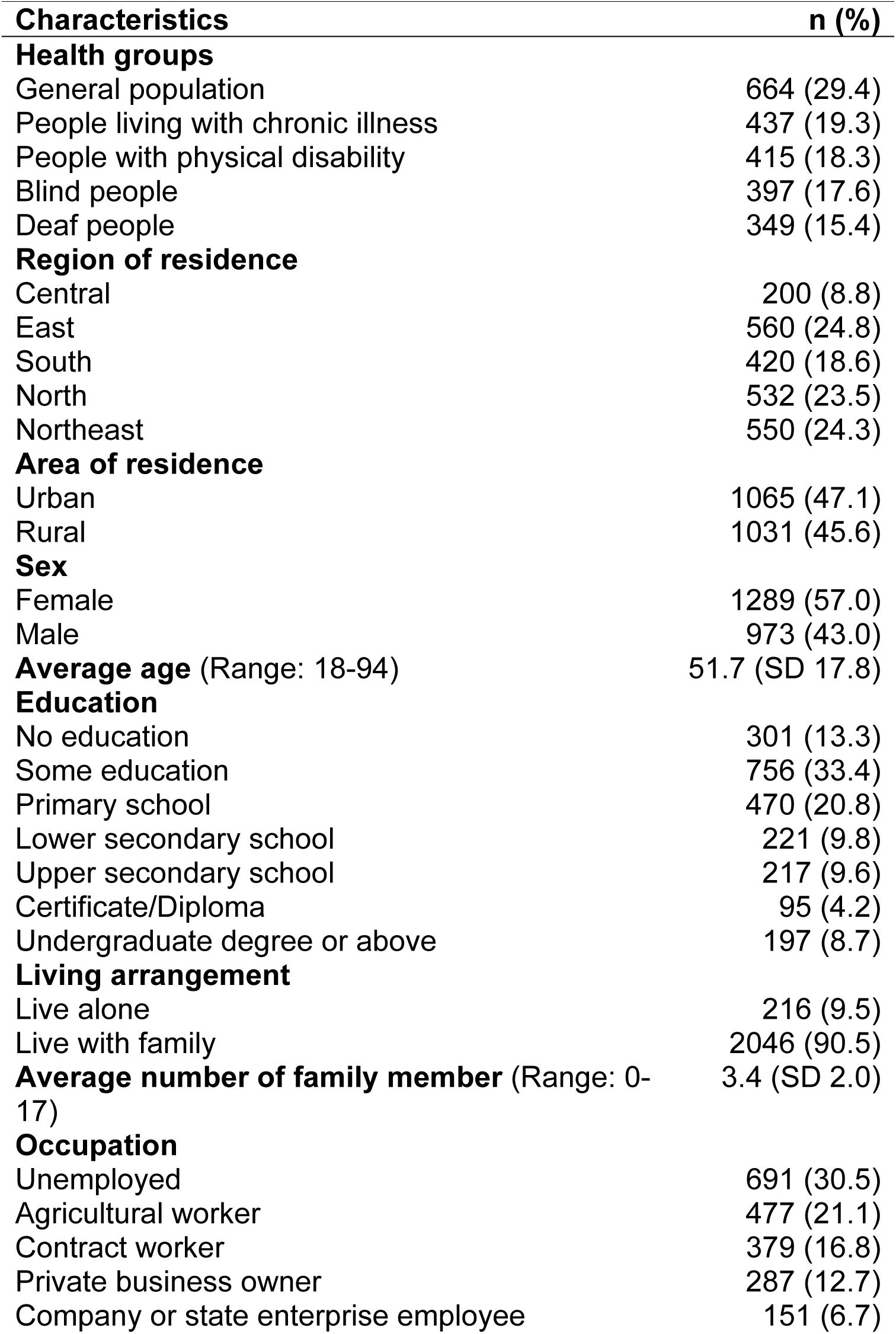
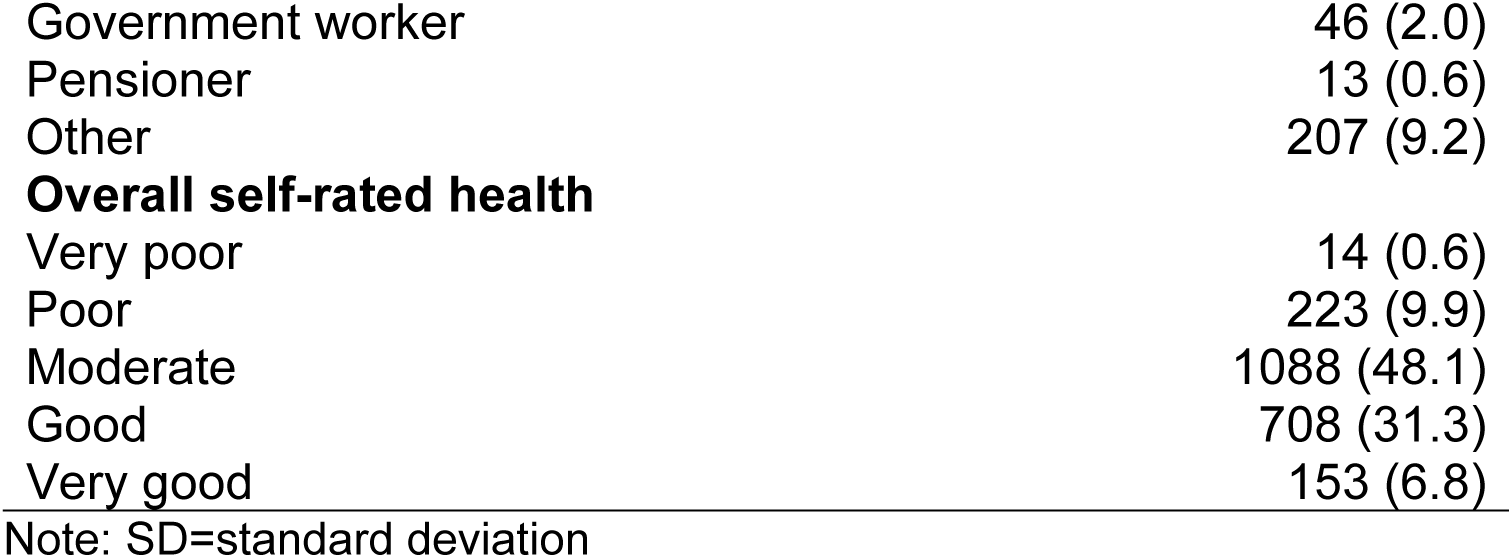
Demographic characteristics of survey participants (N=2,262)

### Health knowledge and capabilities scales

Of the eight Health knowledge and capabilities scales, the one-factor CFA results showed that three scales achieved excellent fit with no statistically significant MIs while one scale had reasonable fit. For the remaining four scales, reasonable to excellent model fits were achieved following model modifications (see Table 3 for a summary and S3 Table for full details of model fit statistics, ranges of factor loadings, relevant confidence intervals and additional results of the alignment analyses).

**Table 3.**
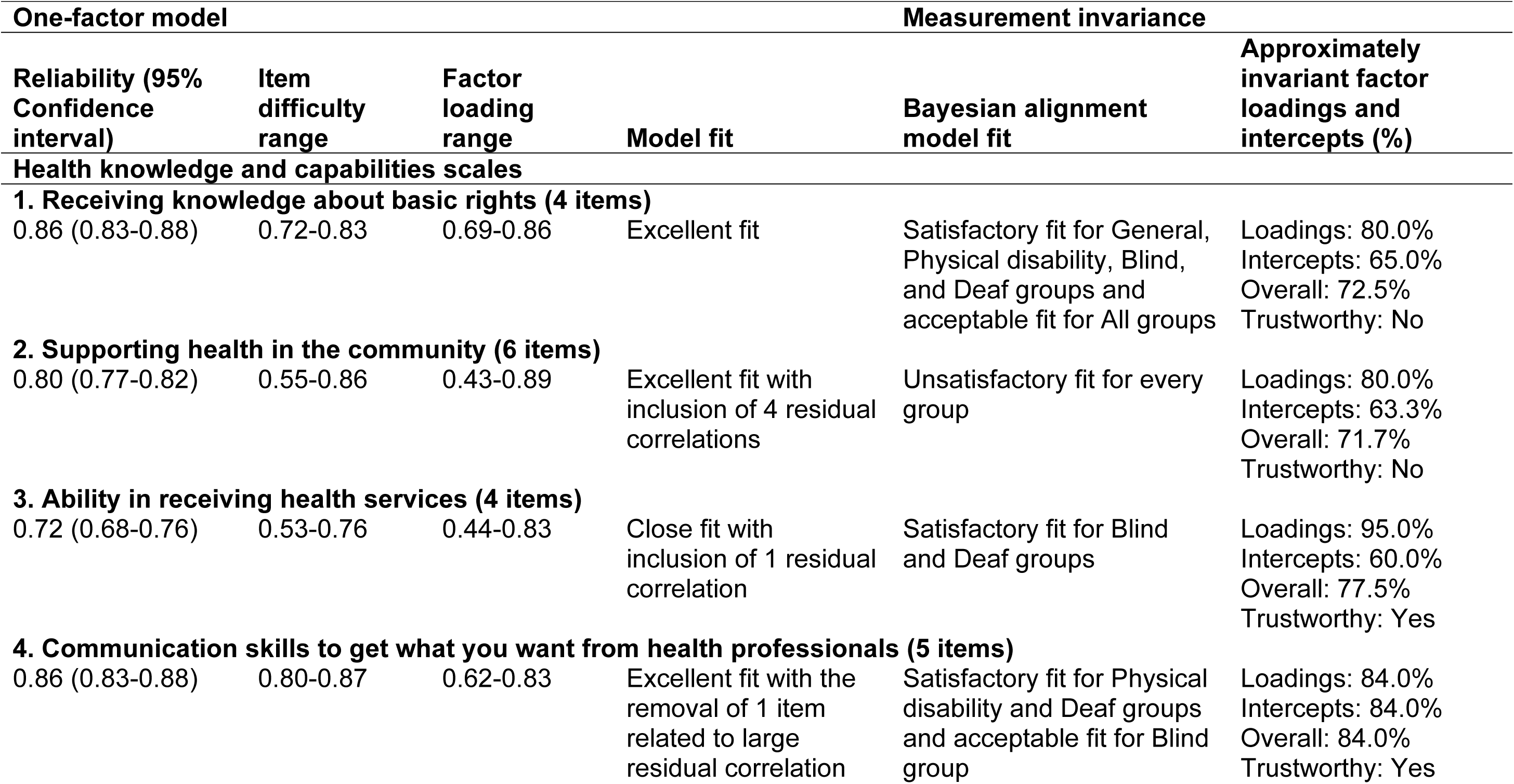
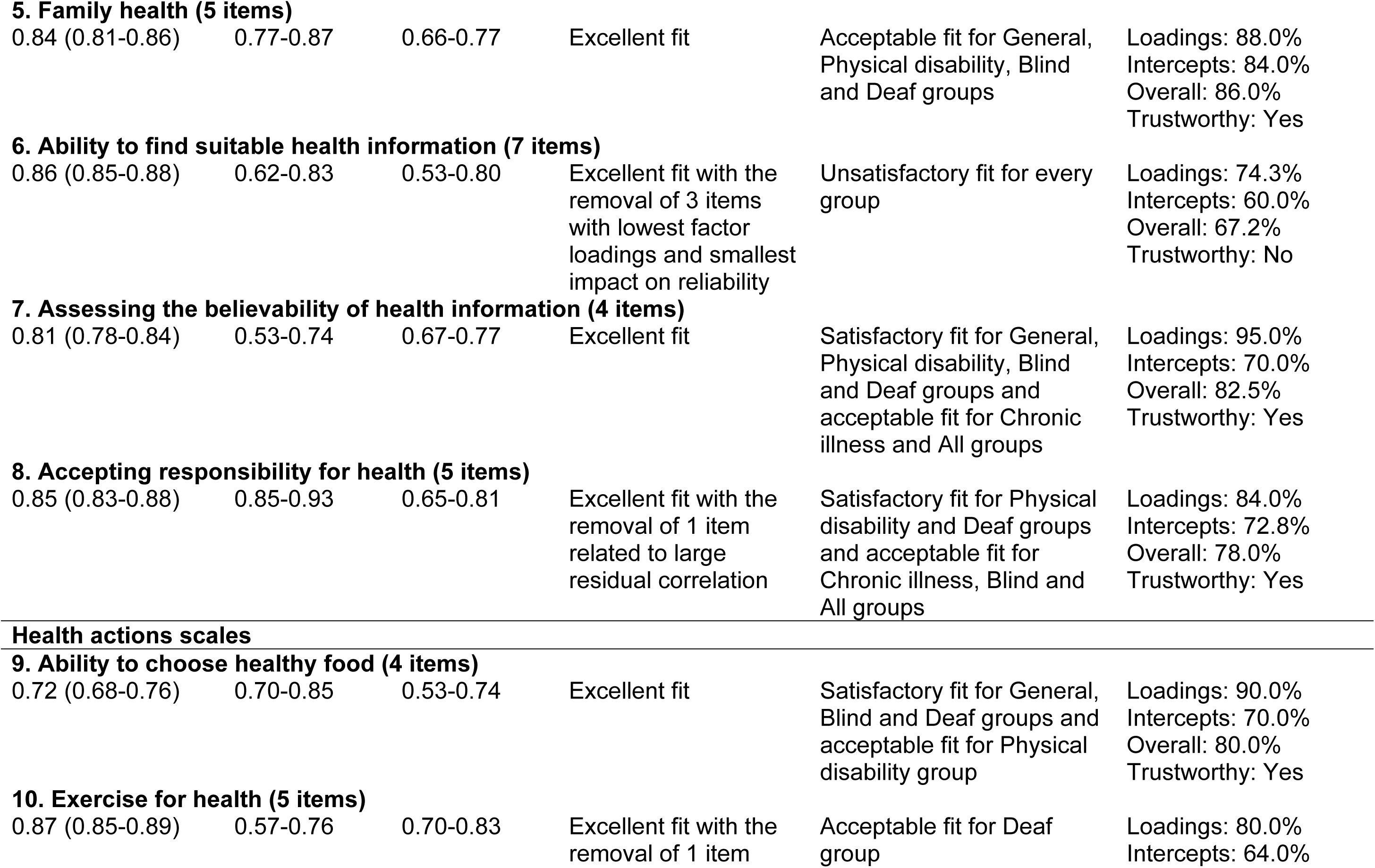
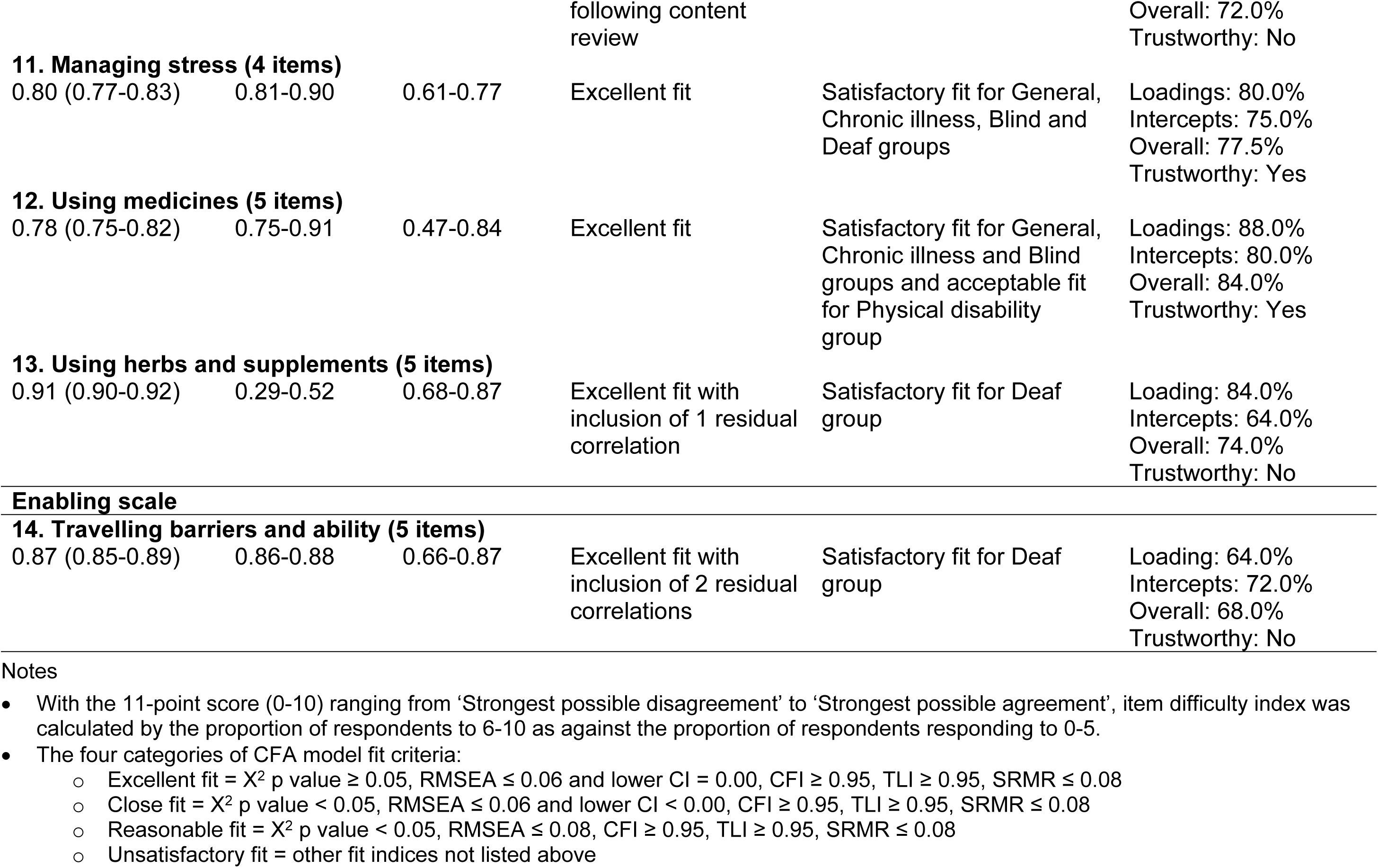
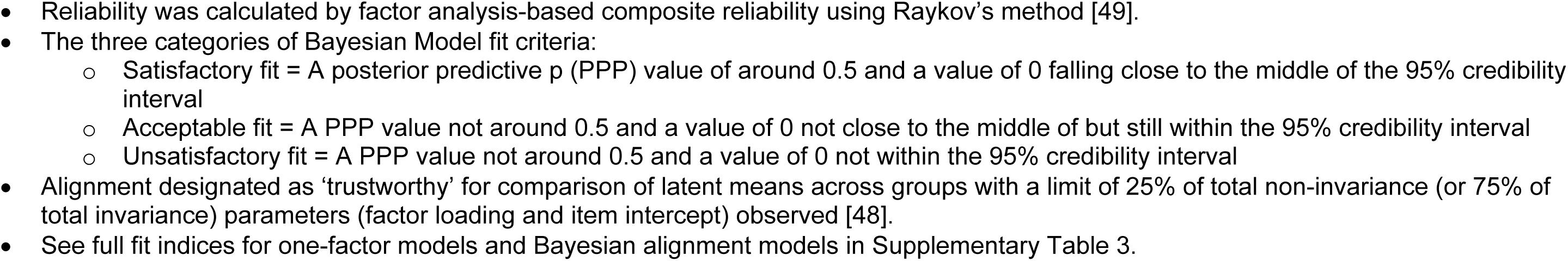
Summary of the psychometric properties and measurement invariance testing of the Information and Support for Health Actions Questionnaire (ISHAQ) with 14 scales and 68 items. One-factor model analyses: General population (n=664) Measurement invariance analyses: General population (n=664), Chronic illness group (n=437), Physical disability group (n=415), Blind group (n=397), Deaf group (n=349), All groups (N=2262)

#### Scale 1. Receiving knowledge about basic rights

Scale 1 had excellent fit with no statistically significant MI in the General Population group. The factor loadings of all items were above 0.60 and the summated scale demonstrated good reliability (0.86). However, the data from the Chronic illness group did not fit the model well and the total invariance (72.5%) led to over the limit of 25% of total non-invariance parameters in the alignment analysis. Nevertheless, the results, combined with content analysis of the items, showed that the items of this scale were acceptably homogeneous, noting that the Chronic illness group might have a more complex concept about receiving knowledge about their rights.

#### Scale 2. Supporting health in the community

Four correlated residuals were identified for the 6-item Scale 2. While one item had a factor loading of only 0.43, a modified model resulted in excellent fit. The pattern of the correlated residuals suggested that a 2-factor solution might be appropriate. This was confirmed by testing a 2-factor ESEM. However, the 2-factor structure did not suggest a clear conceptual difference between the sub-factors. The 6-item model also did not achieve model fit for every group in the alignment analysis while the total invariance was also below the trustworthy threshold (71.7%). Consideration of removing two items related to the correlated residuals led to reasonable fit for the one-factor model and satisfactory fit for the Chronic illness, Physical disability and Deaf groups. However, content analysis showed that one of the removed items was a critical component about health support resources in the neighbourhood. Hence, removal of the item was considered inappropriate. The research team eventually decided to remove only the item with the lowest factor loading.

#### Scale 3. Ability in receiving health services

An item with a low factor loading (0.44) was found for Scale 3 and the initial model fit was unsatisfactory with two statistically significant MIs associated with SEPCs ≥ 0.2. A modified model with a correlated residual (=0.42) produced considerable improvement to close fit. Removal of any of the items with the correlated residual would lead to lower reliability while deletion of the item with the lowest factor loading marginally increased the reliability. The 4-item scale only achieved acceptable fit for the Blind and Deaf groups but did have a total invariance of 77.5%, i.e. alignment results trustworthy. Following discussion among the research team and a review of the cluster analysis results from the concept mapping workshops, it was decided to remove the item with the lowest factor loading and two additional new items were added, making Scale 3 a 5-item scale.

##### Scale 4. Communication skills to get what you want from health professionals

A reasonable fit was found for the one-factor CFA results of Scale 4, with factor loadings ranging from 0.62 to 0.83 and a good reliability of 0.86. However, there was one statistically significant MI associated with SEPC ≥ 0.2. A modified model with a correlated residual (=0.32) led to excellent fit while removal of one of the items with the large correlated residual also achieved excellent fit. The alignment results of the 5-item scale showed trustworthiness (84.0%), but model fit was not achieved for the General population group, the Chronic illness group and All groups. Based on the one-factor CFA results, a 4-item scale was tested and resulted in close to excellent fit for all groups and a total invariance of 90.0%. Hence, this item was removed for the final version.

### Scale 5. Family health

For Scale 5, the 5-item scale had excellent fit for the one-factor CFA, with loadings all above 0.60 and good reliability of 0.84. While the alignment results showed trustworthiness (86.0%), model fit was not achieved for All groups and the Chronic illness group. Further investigation of the Chronic illness group identified a correlated residual (=0.35) between two items. Removal of one of the items led to acceptable or satisfactory fit for every group with trustworthiness being maintained (total invariance of 82.5%). Hence, the recommended item was removed, leading to a revised 4-item scale for Scale 5.

### Scale 6. Ability to find suitable health information

Similar to Scale 2, the ESEM approach was used in the testing of Scale 6. This scale included seven items and the CFA identified seven statistically significant MIs associated with SEPCs ≥ 0.2. Stepwise fitting of correlated residuals led to reasonable fit with the inclusion of five of the recommended residual correlations. Using a 2-factor ESEM analysis and eventual stepwise removal of items with the lowest factor loadings and minimum impact on reliability led to the decision to delete three items, which produced a model with excellent fit. In the invariance analysis, the 7-item model also resulted in unsatisfactory fit for every group while the revised 4-item model fitted the data well for every group except the Physical disability group. The total invariance also increased from 67.2% for the 7-item scale to 80.0% for the 4-item scale.

### Scale 7. Assessing the believability of health information

With four items, the model fit of Scale 7 was excellent with no statistically significant MI. The scale demonstrated a good range of item difficulty (0.53-0.74) with all factor loadings being above 0.60, and good reliability (0.81). This scale also demonstrated satisfactory fit across all special needs groups and a total invariance of 82.5%, indicating that the alignment result was trustworthy.

### Scale 8. Accepting responsibility for health

Finally, the data of the 5-item Scale 8 did not fit the initial model well with one statistically significant MI associated with SEPC ≥ 0.2. A modified model did not improve the fit but yielded another significant MI. Removing one item eventually resulted in excellent fit. The 4-item model also produced satisfactory fit for every group in the alignment analysis, with a total invariance of 97.5%, comparing to the 78.0% of total invariance of the original 5-item model.

## Health actions and enabling scales

Three out of the five health actions scales were found to have excellent fit for their respective hypothesized models. The other two scales achieved close to excellent fit after modifications. The Enabling scale required modification to yield excellent model fit (see Table 3 for a summary and S3 Table for full details).

### Scale 9. Ability to choose healthy food

While two items of Scale 9 had factor loadings below 0.60 but still above 0.50, the model fit was excellent. For the alignment analysis, unsatisfactory fit was found for the Chronic illness group and All groups, but a total invariance of 80.0% was found to be trustworthy. Further investigation could not detect any models with satisfactory fit for the Chronic illness group. Therefore, all items were retained for this scale, but it was annotated that the Chronic illness group might have a sophisticated understanding of the possible differentiation between knowledge and behaviour leading to a possible sub-factor structure for this group.

### Scale 10. Exercise for health

The data fitted to the model of Scale 10 was reasonable while factor loadings were all above 0.60 and reliability was also good (0.87). There were two statistically significant MIs associated with SEPCs ≥ 0.2. Inclusion of one of the suggested correlated residuals led to close fit. Deletion of one item that contributed to a large correlated residual produced an excellent fit with no significant MI. The 4-item model was also tested in the alignment analysis and yielded satisfactory fit for every group, however, the total invariance was only 70%. Nevertheless, the tested item was removed, leading to a 4-item scale.

### Scale 11. Managing stress

Excellent fit was found for Scale 11 with no significant MI. The alignment analysis found that the data of the Physical disability and All groups did not fit the model well while a total invariance of 77.5% was identified. A satisfactory model fit was achieved for the Physical disability group by including a correlated residual (=0.53), suggesting some conceptual overlap among the two items with correlated residuals for this particular group. Content analysis decided that this 4-item scale was homogeneous but might be more complex for people with physical disability. Therefore, all items were retained for Scale 11.

### Scale 12. Using medicines

Scale 12 was another scale with excellent fit. However, this scale had two items with lower factor loadings (0.47 and 0.50) while the reliability was acceptable (0.78). In the alignment analysis, satisfactory fit was achieved for every group except for the Deaf group and All groups, with a total invariance of 84.0%. Removing the item with second lowest factor loading improved the reliability from acceptable to good (0.82) and satisfactory fit could also be achieved by every group. However, this item was considered a critical component of the appropriate use of medicines and removal of this item could diminish the breadth of this scale. Therefore, all items were retained.

### Scale 13. Using herbs and supplements

The initial model fit for the 5-item Scale 13 was not acceptable with two statistically significant MIs associated with SEPCs ≥ 0.2. A modified model with the largest correlated residual (=0.32) led to excellent fit. The 5-item scale only achieved satisfactory fit for the Deaf group. A review of the items found that one item was poorly worded and ambiguous in the Thai language. Removal of this item led to satisfactory fit for every group except the Blind group and All groups, while the total invariance also increased from the threshold of not trustworthy (74.0%) to trustworthy (82.5%). Based on this finding, the poorly worded item was deleted for Scale 13.

### Scale 14. Travelling barriers and ability

The model for Scale 14, the only Enabling scale, did not fit the data well. This scale had the smallest range of item difficulty (0.86-0.88) across all scales but good factor loadings (all above 0.60) and reliability (0.87). Fitting a modified model with two correlated residuals (=0.28 and =0.32) led to good fit. In the alignment analysis, the original 5-item model also did not have satisfactory fit except for the Deaf group with a total invariance of 68.0%. Removing one item related to the correlated residual did not achieve satisfactory fit across every group except acceptable for the Blind group. A content analysis found this scale to be problematic with poor homogeneity and all items were relatively ‘easy’. A final decision was to remove the item with the lowest factor loading (this item was also one of the items with a large correlated residual), leading Scale 14 to be a 4-item scale.

## Supplementary scales for special needs groups

Further testing was undertaken for supplementary scales for special needs groups (see Table 4 for a summary and S4 Table for details). There are two scales for people with chronic illness. The initial models of both scales did not fit well and required modifications to achieve excellent fit. The one-factor model CFA for Scale C1. *Exchange experience and knowledge with other patients* identified two statistically significant MIs associated with SEPCs ≥ 0.2. Including the suggested correlated residuals led to excellent fit. The two pairs of items showed considerable redundancies but removing items from each pair would lead to a 3-item scale. On the other hand, the psychometric data did not provide a clear indication for which item to remove. Following discussion among the research team, the item with the additional knowledge concept was removed. The 4-item model yielded an excellent fit and good reliability (0.88). For Scale C2. *Self-monitoring*, four statistically significant MIs with SEPCs ≥ 0.2 led to incremental fitting of three suggested correlated residuals and improved the fit to close fit. Trial to remove four items was also undertaken, with the 4-item scale resulted in excellent fit and good reliability (0.85). Hence, the final version of Scale C2 included only four items.

**Table 4.**
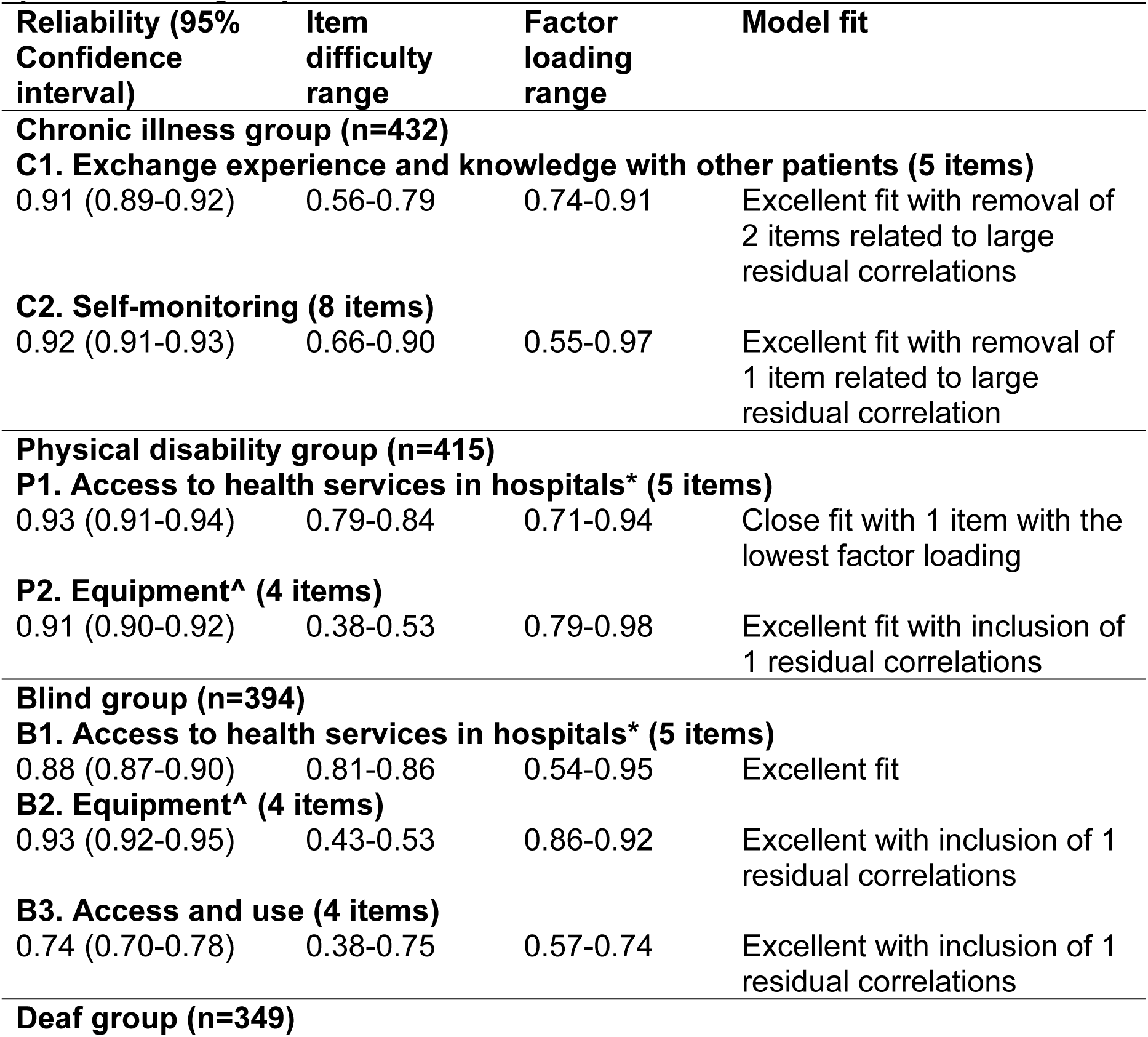
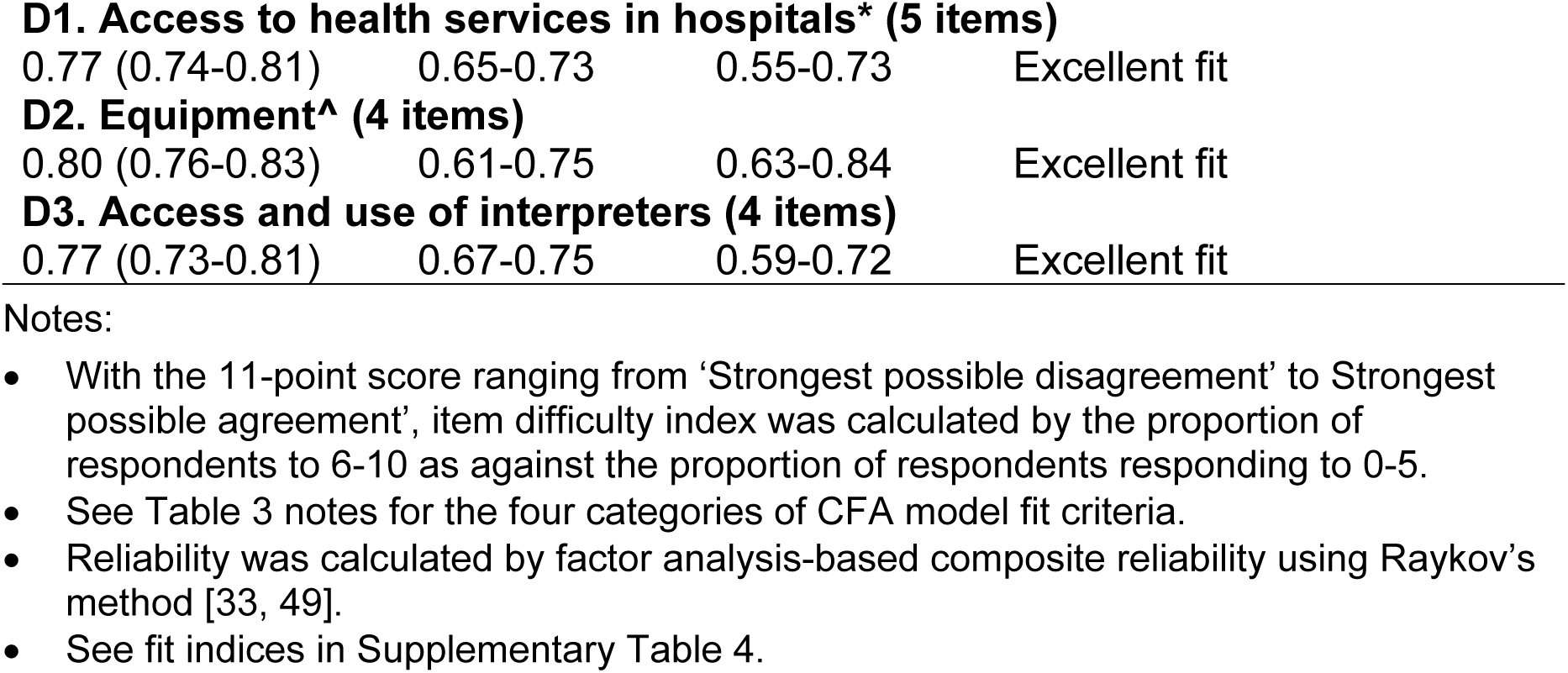
Summary of the psychometric properties of supplementary scales of the Information and Support for Health Actions Questionnaire (ISHAQ) for special needs groups.

Two supplementary scales were developed for people with physical disability, blind and deaf people. Scale P1/B1/D1. *Access to health services in hospitals* did not perform well for the Physical disability group. Initial model fit was not satisfactory with one statistically significant MI associated with SEPC ≥ 0.2. Fitting the suggested correlated residual improved fit to excellent. Consideration was also given to removal of the item with the lowest factor loading and a model with close fit was achieved. However, this scale had excellent fit for the Blind and Deaf groups with no statistically significant MI and reliability was acceptable (Deaf group: 0.77) to good (Blind group: 0.88). Therefore, it was decided that this scale remained as a 5-item scale.

For Scale P2/B2/D2. *Equipment*, initial model fit was not satisfactory for the Physical disability and Blind groups but excellent for the Deaf group. For the Physical disability group, there were two statistically significant MI associated with SEPC ≥ 0.2. Including one or the other of the correlated residual yielded excellent fit. It was the same for the Blind group when including one or the other of the correlated residuals led to excellent fit. However, the data of the Deaf group fit the model well with excellent fit and no statistically significant MI. Reliability was good for all three groups. This scale might not be homogenous for the Physical disability and Blind groups, but it was acceptably homogeneous for the Deaf group. Following discussion among the research team, it was decided that all four items would be retained.

Scale B3. *Access and use* was specifically designed for the Blind group and had four items. Model fit was a close fit, but with one statistically significant MI associated with a SEPC ≥ 0.2. Including the recommended residual correlation yielded an excellent fit. There were no indications from the data that the scale could be improved. Therefore, all items were retained. For the Deaf group, Scale D3. *Access and use of interpreters* fit the data well with no statistically significant MI. This scale was homogeneous with acceptable reliability (0.77) and required no revision.

### Response options

The ISHAQ used a 11-point of a 0-10 scale from ‘strongest possible disagreement’ to ‘strongest possible agreement’. With the 14 main scales, the distribution of all items was significantly negatively skewed, as was the distribution of the total score except for Scale 13. *Using herbs and supplements* where the distribution of one item was significantly positively skewed while the distribution of three items and the total score were significantly positively skewed. This pattern of distribution was also seen in most of the supplementary scales. Combined with experience from the field noting that some participants had problem responding to the 11-point response, the research team recommended changing the response option from a 11-point scale to a 4-point scale of ‘Strongly disagree’, ‘Disagree’, ‘Agree’ and ‘Strongly agree’ with a score of 1 to 4 for each scale.

## Finalised version of the ISHAQ

Following the rigorous psychometric testing, a version of the ISHAQ with eight Health knowledge and capabilities scales and six Health actions scales (with the enabling scale merged with the original five Health actions scales), together with supplementary scales for people with chronic illness (two scales), people with physical disability (two scales), blind people (three scales) and deaf people (three scales) was finalised. Some of the scale names were revised to provide a clearer description of the construct corresponding to the scale. See Table 5 for the ISHAQ scales with high and low descriptors of each construct. See S5 Table for the truncated items of the full ISHAQ (full items are available on request).

**Table 5.**
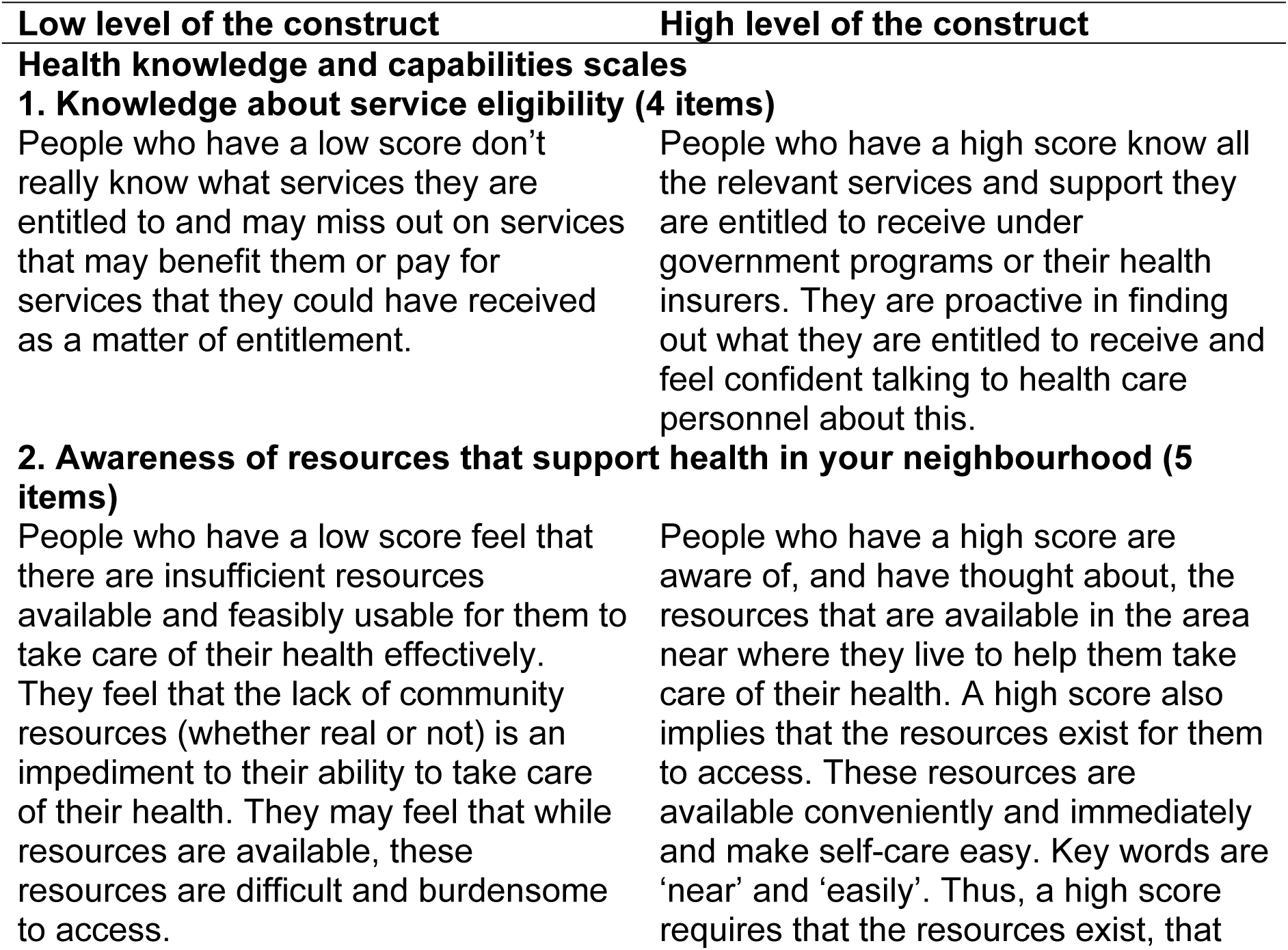
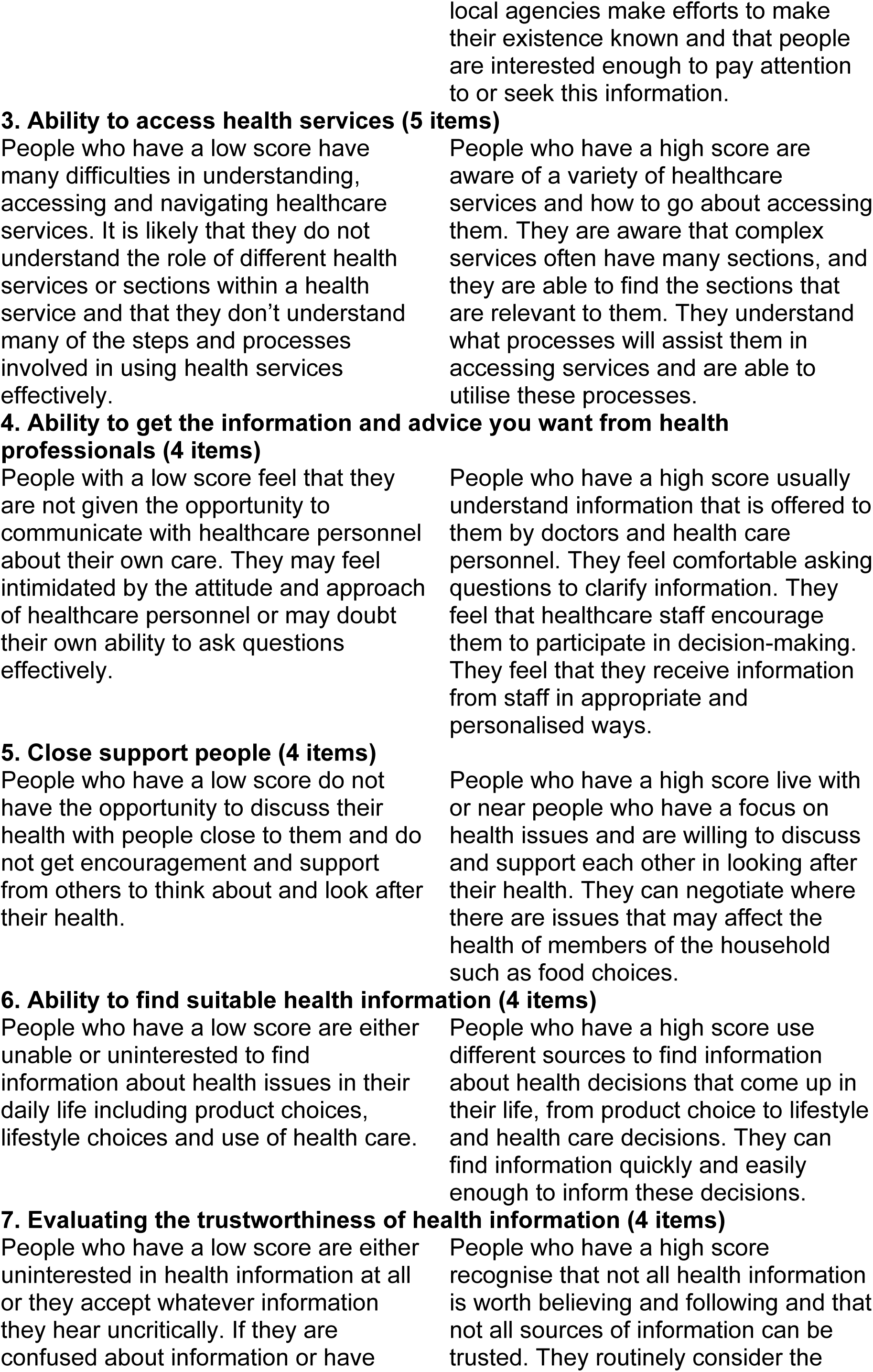
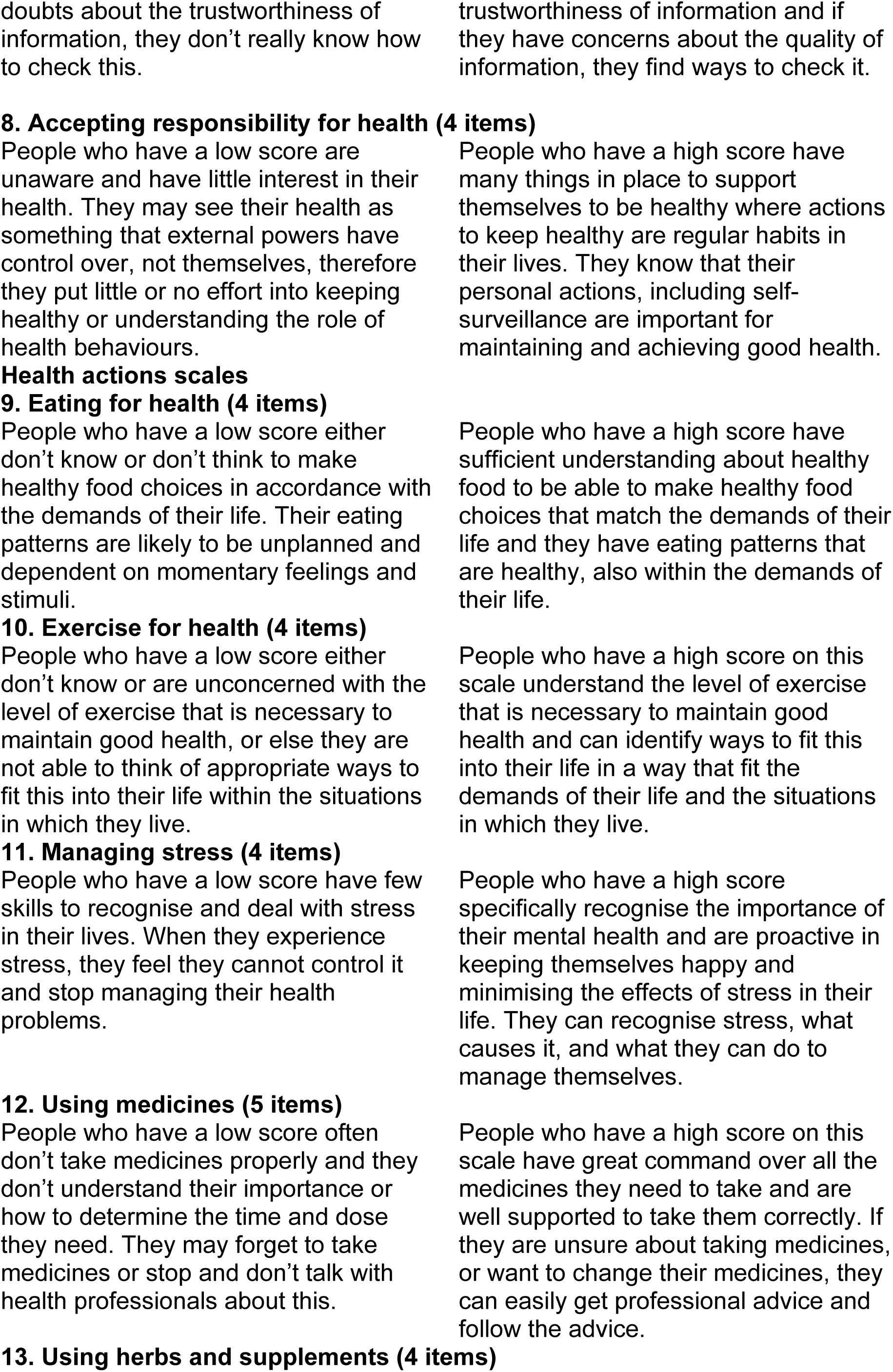
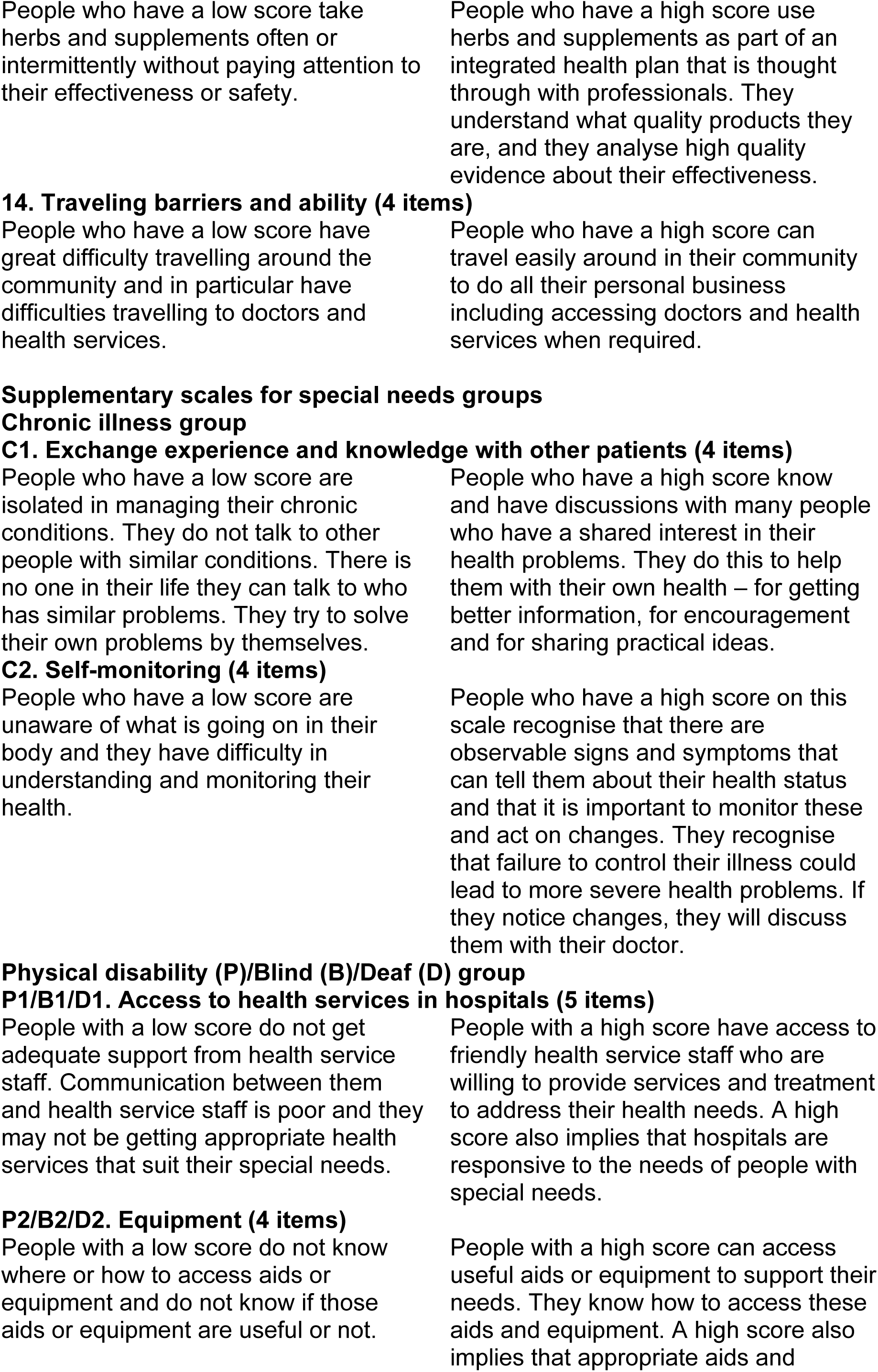
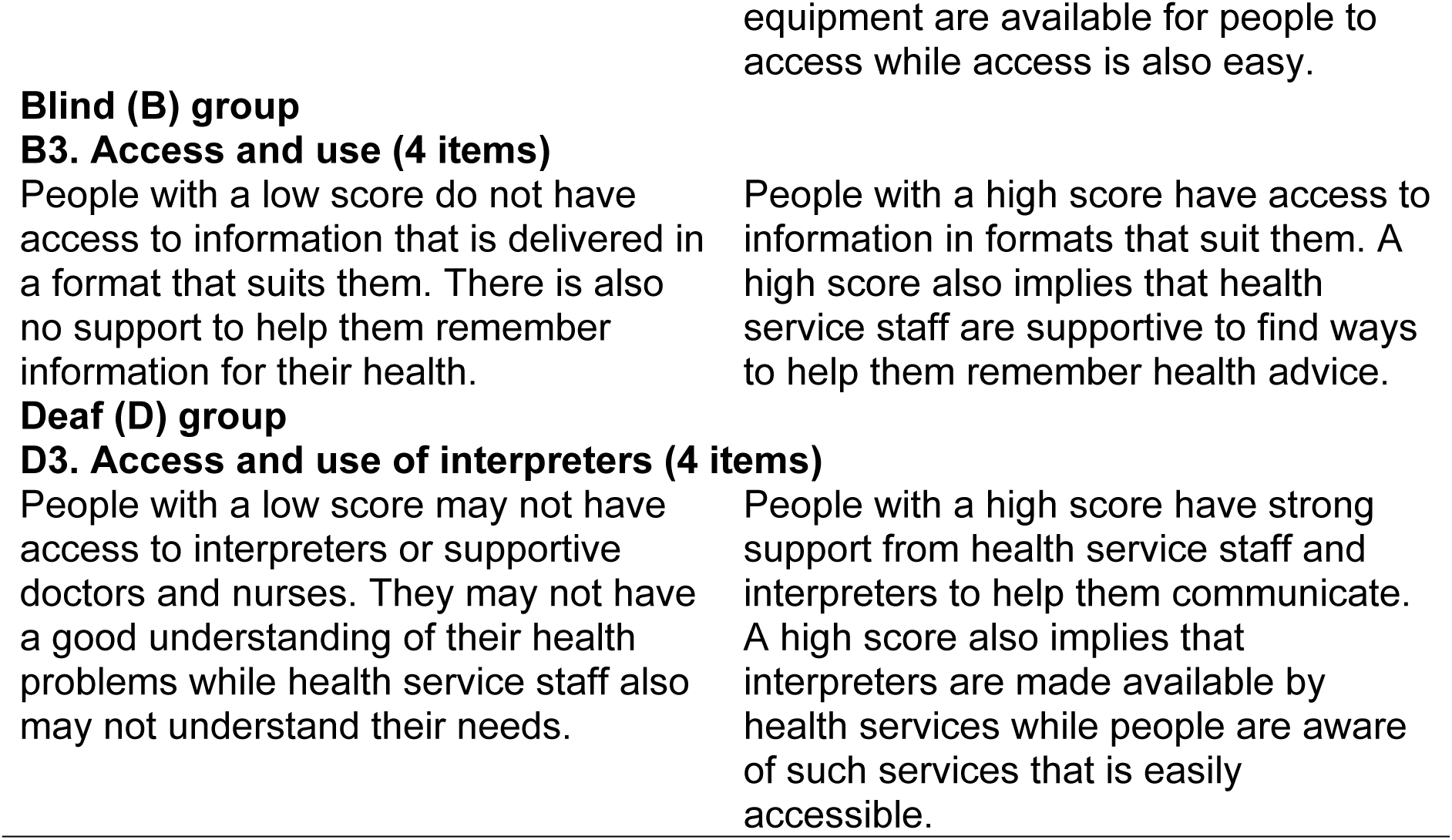
The Information and Support for Health Actions Questionnaire (ISHAQ) scales with high and low descriptors of each construct.

## Discussion

Capturing the different worldview and perspective of the communal culture, a new equitable health literacy measure, the Information and Support for Health Actions Questionnaire (ISHAQ), was conceptualized and developed using a grounded approach in Thailand. This study used a systematic process to test this new health literacy questionnaire and found rigorous validity evidence on the internal structure and reliability of the ISHAQ of the proposed 14 scales as well as the supplementary scales tailored to the needs of people with special health needs. The ISHAQ has now been translated into English and is ready for further validity testing to support the interpretation of its scores as the profiles of people represented by the hypothesised health literacy domains. This questionnaire is expected to be used for health literacy needs assessment, intervention development and evaluation to improve health outcomes and equity.

While it is recommended that the development of health literacy measures should start with a conceptual framework drawn from theory and the literature, which is how most health literacy tools were developed [14], we were aware that the health literacy literature was based on the individualist perspective with an emphasis on individual autonomy and choices. These values might not be applicable in societies where family, community or cultural practices were all intermingled with daily habits, traditions and community behaviours [17]. Drawing from the views of the general population, people with special health needs, health service providers and health policy makers involving 254 participants across 13 concept mapping workshops, followed by an initial validity testing, the ISHAQ conceptualized 14 domains of health literacy and two to three domains for people with special health needs based on the communal perspective. This rigorous process has supported the conceptualisation of the construct and systematic identification of relevant domains and indicators to avoid the pitfall of construct underrepresentation [5]. The only commonly used health literacy instrument that also used a grounded approach through extensive consultation is the HLQ, which identified nine dimension of health literacy [16].

Comparing to the HLQ, there are similarities around the health literacy domains that link to health information, healthcare providers, health services and social support. However, the ISHAQ also covers the domain of resources in the neighbourhood and more practical domains about eating, exercising, managing stress, using medicines, herbs and supplements as well as about the enabling factor of travelling for health actions. The scales for people with chronic illness also include a scale about interaction with other people to share and exchange experiences and knowledge and self-monitoring. These domains are tapping into the daily life and interactions between people, community, health services and their environment. Using the ISHAQ as a health literacy needs assessment tool will provide a holistic picture of where people’s strengths or challenges are in managing their health. It will also imply if their health services, healthcare providers and surrounding environment are responsive to or supportive of their health needs.

This study used a rigorous and systematic approach to test for the validity evidence of the ISHAQ and help refined the tool for further validity testing and dissemination. A series of one-factor models using data from the general population as the reference group was first undertaken. All scales demonstrated acceptable to good reliability, while 11 out of the 14 scales had composite reliability of ≥ 0.80. The reliability of Scales 3, 9 and 12 were a little lower but were all above 0.70, which is still within the acceptable range. Eight scales (Scales 1, 4, 5, 8, 9, 11, 12 and 14) had quite a restrictive range of item difficulty (0.02-0.16) and these scales had a median ‘difficulty’ (in the direction of ‘easiness’) of 0.70 or higher. The remaining scales (Scales 2, 3, 6, 7, 10 and 13) showed a good range of difficulty, indicating the tool’s sensitivity to identify people with different levels of capability within the relevant domain.

Given the scales were set a priori, CFAs were conducted for each of the scales. Of the 14 scales, six scales achieved excellent fit with no statistically significant MI associated with SEPC ≥ 0.2. By using residual correlations recommended by the modification indices for model modifications, all other scales yield reasonable to excellent fits. The modification results, combined by content analysis, was then used to informed further testing, using a different approach such as ESEM if needed to identify sub-structure or removing conceptually redundant items.

With the aim to develop an equitable measure, measurement invariance testing across different special needs groups using the alignment method was also undertaken. Based on the threshold proposed by Asparouhov and Muthén [48], the developers of this method, the results indicated that trustworthy alignment could be established for eight of the 14 scales, while the trustworthiness of six scales could not be ascertained. The results seemed to point more to the issue of noninvariant intercepts (scalar invariance) for the six scales. Model fit also could not be established for every group for most of the scales. These results were then used to inform and support revision of the scales.

One-factor CFAs were also undertaken for the supplementary scales for the Chronic illness, Physical disability, Blind and Deaf groups. Interestingly, the three scales for the Deaf group all yield excellent fit, with acceptable to good reliability. The two scales of the Chronic illness group required model modifications due to correlated residuals, same as the two scales of the Physical disability group. Two of the three scales for the Blind group had excellent fit while one required modification.

Following this rigorous validity testing process, a revised version of the ISHAQ with 14 scales (59 items) and supplementary scales for special needs groups (two scales with for the Chronic illness group, two scales for the Physical disability group, three scales for the Blind group and three scales for the Deaf group) was generated. Initial testing of six of the revised models (Scales 4, 5, 6, 8, 10 and 13) achieved reasonable to excellent model fit for the general population group as well as leading to trustworthy measurement invariance results. Scale 3. *Ability in receiving health services* was not tested due to the decision to add two new items. The two revised scales for the Chronic illness group also achieved excellent fit. Considered that validity testing is an ongoing process [5], further testing of the tool is warranted to contribute to the validity evidence accumulated.

Although the ISHAQ was developed in Thailand which is a predominate communal culture, it is important to note that cultures exist in a continuum where there are always varying degrees of communal and individualistic influences within a society or even within a certain context or setting [17, 50]. For example, communal cultures are often evident in the rural or regional communities in America or England [51, 52]. Therefore, the ISHAQ also has the potential to be applied in other countries. The 14 constructs can just be as relevant to people living in a predominately individualistic culture. For example, in a 2021-2022 survey, 49.4% of Australians reported using herbs or supplements over the previous 12 months [53], showing that Scale 13. *Using herbs and supplements* is not unique to people coming from a communal culture with the preference to use traditional medicines. Transportation, as measured by Scale 14. *Traveling barriers and ability*, has also been found to be a barrier for accessing healthcare in the United States either in rural or urban areas [54]. With the experience from the Covid-19 pandemic, the domains measured by Scales C1. *Exchange experience and knowledge with other patients* and C2. *Self-monitoring* for people living with chronic illness may just as applicable for people experiencing a non-chronic condition. Hence, these two scales also have the potential to be adapted to the general population to understand their behaviour when facing certain health conditions.

With the ISHAQ ready to be used in different countries and settings, it is also important to note that the ISHAQ, being a multi-dimensional tool, uses scale scores instead of an overall sum score for its results. As such, healthcare providers and policy makers using this tool can gain insight into the health literacy patterns of their group of interests. It will allow for the use of a strengths-based approach to develop interventions that build on health literacy strengths (scales with higher scores) to support challenges (scales with lower scores) instead of just taking the usual deficit-based approach of classifying people into high/adequate or low/inadequate health literacy [4]. It is expected that a survey using the ISHAQ will have the potential to support health practitioners and policy makers to improve people’s health outcomes and equity.

## Strengths and Limitations

This study used a grounded approach involving 254 people including the general population, people with special health needs, healthcare professionals and policy makers to conceptualise health literacy based on real-life experiences. By using the most sophisticated psychometric testing methods available, the ISHAQ, with strong validity evidence of internal structure and reliability, was developed and ready for further testing and use.

It should be noted that we did not conduct a final multi-factor analysis as recommended in the usual classical factor analytic approach because most of the scales were revised while Scale 3 had recommended items added. Therefore, discriminant validity was not established. Future testing of this finalized ISHAQ version can consider doing multi-factor analysis to confirm the hypothesised structure of the full measurement model as well as investigation of discriminant validity among the health literacy domains. However, given that the ISHAQ was developed based on the communal perspective which views the world as being interconnected instead of self-sufficiency as in the individualist perspective [55], it is likely that the hypothesised dimensions of ISHAQ are also interconnected to a certain degree.

While measurement invariance was conducted for people with chronic illness, people with physical disability, blind people and deaf people, other groups as defined by demographic factors such as age, gender, education were not tested. It is important that measurement invariance among different demographic groups also be established for an equitable measure.

## Conclusion

In identifying a gap in current health literacy measures that support the perspective of people from the communal culture, we used a grounded approach to conceptualise health literacy and subsequently developed the ISHAQ based on people’s real-life experiences. The identified health literacy domains capture how people deal with their health in their daily lives. While the conceptualisation came from a communal perspective, it is obvious that these health literacy domains are also recognisable in societies dominated by individualistic culture. The ISHAQ has opened the opportunity to gain new and deeper insights into people’s health literacy profiles. It can then be used to inform healthcare providers and policy makers to develop interventions that optimise health literacy, leading to improved health outcomes and equity.

## Data Availability

The dataset generated and analysed are not publicly available due to ethical requirement and privacy concern but may be available from the corresponding author on reasonable request.

## Acknowledgments

The authors would like to thank Mrs Budsaraporn Bhechrung from the Health Systems Research Institute for co-ordinating and communicating with local community self-help groups and local authorities. We would also like to thank The National Association of Deaf of Thailand, The Association of the Physically Handicapped of Thailand for their assistance in data collection. We express our gratitude to all our participants who joined the concept mapping workshops and participated in the surveys.

## Author Contributions

**Conceptualization:** Roy Batterham, Gerald R Elsworth, Saichon Kloyiam, Charay Vicathai, Napaporn Wanitkun, Richard H Osborne

**Formal analysis:** Christina Cheng, Roy Batterham, Gerald R Elsworth, Richard H Osborne

**Methodology:** Roy Batterham, Gerald R Elsworth, Richard H Osborne

**Project administration:** Roy Batterham, Saichon Kloyiam, Charay Vicathai , Napaporn Wanitkun

**Writing – original draft preparation:** Christna Cheng

**Writing – review and editing:** Christina Cheng, Roy Batterham, Gerald R Elsworth, Saichon Kloyiam, Charay Vicathai, Napaporn Wanitkun, Melanie Hawkins, Richard H Osborne

## Supporting information

S1 Figure. ISHAQ original measurement model

S2 Table. Study 1 psychometric properties of the 16 one-factor models of the Information and Support for Health Actions Questionnaire (ISHAQ) (N = 2,282)

S3 Table. Psychometric properties and measurement invariance testing of the Information and Support for Health Actions Questionnaire (ISHAQ) with 14 scales and 68 items

S4 Table. Psychometric properties of additional scales of the Information and Support for Health Actions Questionnaire (ISHAQ) for special needs groups S5 Table. Truncated items of the Information and Support for Health Actions Questionnaire (ISHAQ)

## References

1. Paasche-Orlow MK, Wolf MS. Promoting health literacy research to reduce health disparities. J Health Commun. 2010;15 Suppl 2:34–41. Epub 2010/09/29. doi: 10.1080/10810730.2010.499994. PubMed PMID: 20845191.

2. Institute of Medicine Committee on Health Literacy. Health Literacy: A Prescription to End Confusion Nielsen-Bohlman L, Panzer AM, Kindig DA, editors. Washington (DC): National Academies Press (US) Copyright 2004 by the National Academy of Sciences. All rights reserved.; 2004.

3. Batterham RW, Hawkins M, Collins PA, Buchbinder R, Osborne RH. Health literacy: applying current concepts to improve health services and reduce health inequalities. Public Health. 2016;132:3–12. doi: 10.1016/j.puhe.2016.01.001. PubMed PMID: 26872738.

4. Osborne RH, Cheng CC, Nolte S, Elmer S, Besancon S, Budhathoki SS, et al. Health literacy measurement: embracing diversity in a strengths-based approach to promote health and equity, and avoid epistemic injustice. BMJ Global Health. 2022;7(9):e009623. doi: 10.1136/bmjgh-2022-009623.

5. American Educational Research Association (AERA), American Psychological Association (APA), National Council on Measurement in Education (NCME). Standards for educational and psychological testing. Washington, DC: American Educational Research Association; 2014.

6. Simonds SK. Health Education as Social Policy. Health education monographs. 1974;2(1_suppl):1-10. doi: 10.1177/10901981740020S102.

7. Sørensen K, Van den Broucke S, Fullam J, Doyle G, Pelikan J, Slonska Z, et al. Health literacy and public health: A systematic review and integration of definitions and models. BMC Public Health. 2012;12(1):80. doi: 10.1186/1471-2458-12-80.

8. Nutbeam D. Health literacy as a public health goal: a challenge for contemporary health education and communication strategies into the 21st century. Health Promotion International. 2000;15(3):259–67. doi: 10.1093/heapro/15.3.259.

9. Paasche-Orlow MK, Wolf MS. Evidence does not support clinical screening of literacy. J Gen Intern Med. 2008;23(1):100–2. Epub 2007/11/10. doi: 10.1007/s11606-007-0447-2. PubMed PMID: 17992564; PubMed Central PMCID: PMCPMC2173929.

10. Kickbusch I, Maag D, Wait S, Health Af, Future t, UK. ILC. Navigating Health: The Role of Health Literacy: Alliance for Health and the Future, International Longevity Centre-UK; 2005.

11. Berkman ND, Davis TC, McCormack L. Health literacy: What Is It? Journal of health communication. 2010;15((Supp 2)):9-19. doi: 10.1080/10810730.2010.499985.

12. Nutbeam D. The evolving concept of health literacy. Social Science & Medicine. 2008;67(12):2072–8. doi: 10.1016/j.socscimed.2008.09.050.

13. Levin-Zamir D, Leung AYM, Dodson S, Rowlands G. Health literacy in selected populations: Individuals, families, and communities from the international and cultural perspective. Information Services & Use. 2017;37:131–51. doi: 10.3233/ISU-170834.

14. Tavousi M, Mohammadi S, Sadighi J, Zarei F, Kermani RM, Rostami R, et al. Measuring health literacy: A systematic review and bibliometric analysis of instruments from 1993 to 2021. PloS one. 2022;17(7):e0271524. doi: 10.1371/journal.pone.0271524.

15. Sørensen K, Van den Broucke S, Pelikan JM, Fullam J, Doyle G, Slonska Z, et al. Measuring health literacy in populations: illuminating the design and development process of the European Health Literacy Survey Questionnaire (HLS-EU-Q). BMC Public Health. 2013;13(1):948. doi: 10.1186/1471-2458-13-948.

16. Osborne RH, Batterham RW, Elsworth GR, Hawkins M, Buchbinder R. The grounded psychometric development and initial validation of the Health Literacy Questionnaire (HLQ). BMC Public Health. 2013;13(1):658. doi: 10.1186/1471-2458-13-658.

17. World Health Organization. Health literacy development for the prevention and control of noncommunicable diseases. Geneva: World Health Organization, 2022.

18. Bhakuni H, Abimbola S. Epistemic injustice in academic global health. The Lancet Global Health. 2021;9(10):e1465–e70. doi: 10.1016/S2214-109X(21)00301-6.

19. Aaby A, Friis K, Christensen B, Rowlands G, Maindal HT. Health literacy is associated with health behaviour and self-reported health: A large population-based study in individuals with cardiovascular disease. European Journal of Preventive Cardiology. 2017;24(17):1880–8. doi: 10.1177/2047487317729538.

20. Anwar WA, Mostafa NS, Hakim SA, Sos DG, Abozaid DA, Osborne RH. Health literacy strengths and limitations among rural fishing communities in Egypt using the Health Literacy Questionnaire (HLQ). PloS one. 2020;15(7):e0235550. doi: 10.1371/journal.pone.0235550.

21. Gill S, Zeki R, Kaye S, Zingirlis P, Archer V, Lewandowski A, et al. Health literacy strengths and challenges of people in New South Wales prisons: a cross-sectional survey using the Health Literacy Questionnaire (HLQ). BMC Public Health. 2023;23(1):1520. doi: 10.1186/s12889-023-16464-3.

22. Maia AC, Marques MJ, Goes AR, Gama A, Osborne Ri, Dias S. Health literacy strengths and needs among migrant communities from Portuguese-speaking African countries in Portugal: a cross-sectional study. Frontiers in public health. 2024;12. doi: 10.3389/fpubh.2024.1415588.

23. Passi R, Kaur M, Lakshmi PVM, Cheng C, Hawkins M, Osborne RH. Health literacy strengths and challenges among residents of a resource-poor village in rural India: Epidemiological and cluster analyses. PLOS Global Public Health. 2023;3(2):e0001595. doi: 10.1371/journal.pgph.0001595.

24. Beauchamp A, Batterham RW, Dodson S, Astbury B, Elsworth GR, McPhee C, et al. Systematic development and implementation of interventions to OPtimise Health Literacy and Access (Ophelia). BMC Public Health. 2017;17(1):230. doi: 10.1186/s12889-017-4147-5.

25. Borge CR, Larsen MH, Osborne RH, Aas E, Kolle IT, Reinertsen R, et al. Impacts of a health literacy-informed intervention in people with chronic obstructive pulmonary disease (COPD) on hospitalization, health literacy, self-management, quality of life, and health costs – A randomized controlled trial. Patient Education and Counseling. 2024;123:108220. doi: 10.1016/j.pec.2024.108220.

26. Himkhun L, Danaidutsadeekul S, Wanitkun N, Chawasiri C. Predictors of Quality of life among Post-Lumbar Surgery Patients: ปัจจัยทำนายคุณภาพชีวิตผู้ป่วยหลังผ่าตัดกระดูกสันหลังระดับเอว. Nursing Science Journal of Thailand. 2017;35(3):82–93.

27. IBM Corp. IBM SPSS Statistics for Windows. 22.0 ed. Armonk, NY: IBM Corp.; 2013.

28. Muthén LK, Muthén BO. Mplus User’s Guide. Los Angeles, CA: Muthén & Muthén; 2013.

29. Wolf E, Harrington K, Clark S, Miller M. Sample Size Requirements for Structural Equation Models. Educational and Psychological Measurement. 2013;73(6):913–34. doi: 10.1177/0013164413495237.

30. Lord FM, Novick MR, Birnbaum A. Statistical theories of mental test scores. Oxford, England: Addison-Wesley; 1968.

31. Raykov T. Bias of Coefficient afor Fixed Congeneric Measures with Correlated Errors. Applied Psychological Measurement. 2001;25(1):69–76. doi: 10.1177/01466216010251005.

32. Raykov T. Reliability if deleted, not ‘alpha if deleted’: Evaluation of scale reliability following component deletion. British Journal of Mathematical and Statistical Psychology. 2007;60(2):201–16. doi: 10.1348/000711006X115954.

33. Raykov T. Scale construction and development using structural equation modeling. Handbook of structural equation modeling. New York, NY, US: The Guilford Press; 2012. p. 472–92.

34. Sarstedt M, Ringle CM, Hair JF. Partial Least Squares Structural Equation Modeling. In: Homburg C, Klarmann M, Vomberg A, editors. Handbook of Market Research. Cham: Springer International Publishing; 2022. p. 587-632.

35. Hattie JA. Methodology review: Assessing unidimensionality of tests and items. Applied Psychological Measurement. 1985;9(2):139–64. doi: 10.1177/014662168500900204.

36. Ziegler M, Hagemann D. Testing the unidimensionality of items: Pitfalls and loopholes. European Journal of Psychological Assessment. 2015;31(4):231–7. doi: 10.1027/1015-5759/a000309.

37. Tabachnick BG, Fidell LS. Using multivariate statistics, 5th ed. Boston, MA: Allyn & Bacon/Pearson Education; 2007. xxvii, 980-xxvii, p.

38. Dominguez-Lara S. Proposal for cut-offs for factor loadings: A construct reliability perspective. Enferm Clin (Engl Ed). 2018;28(6):401–2. Epub 20180720. doi: 10.1016/j.enfcli.2018.06.002. PubMed PMID: 30037488.

39. Hu Lt, and Bentler PM. Cutoff criteria for fit indexes in covariance structure analysis: Conventional criteria versus new alternatives. Structural Equation Modeling: A Multidisciplinary Journal. 1999;6(1):1–55. doi: 10.1080/10705519909540118.

40. Saris WE, Albert S, and van der Veld WM. Testing Structural Equation Models or Detection of Misspecifications? Structural Equation Modeling: A Multidisciplinary Journal. 2009;16(4):561–82. doi: 10.1080/10705510903203433.

41. Whittaker TA. Using the Modification Index and Standardized Expected Parameter Change for Model Modification. The Journal of Experimental Education. 2012;80(1):26–44. doi: 10.1080/00220973.2010.531299.

42. Marsh HW, Morin AJ, Parker PD, Kaur G. Exploratory structural equation modeling: an integration of the best features of exploratory and confirmatory factor analysis. Annu Rev Clin Psychol. 2014;10:85–110. Epub 2013/12/10. doi: 10.1146/annurev-clinpsy-032813-153700. PubMed PMID: 24313568.

43. Prokofieva M, Zarate D, Parker A, Palikara O, Stavropoulos V. Exploratory structural equation modeling: a streamlined step by step approach using the R Project software. BMC Psychiatry. 2023;23(1):546. doi: 10.1186/s12888-023-05028-9.

44. Millsap RE, Olivera-Aguilar M. Investigating measurement invariance using confirmatory factor analysis. Handbook of structural equation modeling. New York, NY, US: The Guilford Press; 2012. p. 380-92.

45. Muthén BO, Asparouhov T. New methods for the study of measurement invariance with many groups: Statmodel; 2013 [cited 2025 6 February]. Available from: https://www.statmodel.com/download/PolAn.pdf.

46. Luong R, Flake JK. Measurement invariance testing using confirmatory factor analysis and alignment optimization: A tutorial for transparent analysis planning and reporting. Psychological Methods. 2023;28(4):905–24. doi: 10.1037/met0000441.

47. Muthén B, Asparouhov T. Bayesian structural equation modeling: a more flexible representation of substantive theory. Psychol Methods. 2012;17(3):313–35. doi: 10.1037/a0026802. PubMed PMID: 22962886.

48. Muthén B, Asparouhov T. IRT studies of many groups: the alignment method. Frontiers in Psychology. 2014;5. doi: 10.3389/fpsyg.2014.00978.

49. Raykov T, Marcoulides GA. Introduction to psychometric theory. New York, NY, US: Routledge/Taylor & Francis Group; 2011. xii, 335-xii, p.

50. Green EGT, Deschamps J-C, Páez D. Variation of Individualism and Collectivism within and between 20 Countries: A Typological Analysis. Journal of Cross-Cultural Psychology. 2005;36(3):321–39. doi: 10.1177/0022022104273654.

51. Riney-Kehrberg P. The Routledge history of rural American: Routledge; 2016.

52. Neal S, Walters S. Rural Be/longing and Rural Social Organizations: Conviviality and Community-Making in the English Countryside. Sociology. 2008;42(2):279–97. doi: 10.1177/0038038507087354.

53. Harnett J, McIntyre E, Adams J, Addison T, Bannerman H, Egelton L, et al. Prevalence and Characteristics of Australians Complementary Medicine Product Use, and Concurrent Use with Prescription and Over-the-Counter Medications-A Cross Sectional Study. Nutrients. 2023;15(2). Epub 20230109. doi: 10.3390/nu15020327. PubMed PMID: 36678198; PubMed Central PMCID: PMCPMC9860983.

54. Syed ST, Gerber BS, Sharp LK. Traveling Towards Disease: Transportation Barriers to Health Care Access. Journal of community health. 2013;38(5):976–93. doi: 10.1007/s10900-013-9681-1.

55. Triandis HC. Individualism & collectivism. Individualism & collectivism: Westview Press; 1995. p. xv, 259-xv, .

